# Discovering Genetic Signatures Associated with Alzheimer’s Disease in Tiled Whole Genome Sequence Data: Results from the Artificial Intelligence for Alzheimer’s Disease (AI4AD) Consortium

**DOI:** 10.1101/2024.08.01.24311329

**Authors:** Sarah Wait Zaranek, Alexander Wait Zaranek, Peter Amstutz, Jingxuan Bao, Jiong Chen, Tom Clegg, Hannah Craft, Taeho Jo, Brian Lee, Kwangsik Nho, Sophia I. Thomopoulos, Christos Davatzikos, Li Shen, Heng Huang, Paul M. Thompson, Andrew J. Saykin, the Alzheimer’s Disease Neuroimaging Initiative (as a consortium author) for the AI4AD Initiative

**Author notes:** These authors contributed equally (Sarah Wait Zaranek, Alexander Wait Zaranek, Christos Davatzikos, Li Shen, Heng Huang, Paul M. Thompson, Andrew J. Saykin). Submitted for peer review Pacific Symposium on Biocomputing (PSB) 2025.

## Abstract

Currently, the ability to analyze large-scale whole genome sequence (WGS) data is limited due to both the size of the data and the inability of many existing tools to scale. To address this challenge, we use data “tiling” to efficiently partition whole genome sequences into smaller segments resulting in a simple numeric matrix of small integers. This lossless representation is particularly suitable for machine learning (ML) models. As an example of the benefits of tiling, we showcase results from tiled data as part of the Artificial Intelligence for Alzheimer’s Disease (AI4AD) consortium. AI4AD is a coordinated initiative to develop transformative AI approaches for high throughput analysis of next generation sequencing and related imaging, AD biomarker, and cognitive data. The collective effort integrates imaging, genomic, biomarker, and cognitive data to address fundamental barriers in AD prevention and drug discovery. One of the project’s initial aims is to discover new genetic signatures in WGS data that can be used to understand AD risk and progression in conjunction with imaging, biomarker and cognitive data. We tiled and analyzed 15,000+ genomes from the Alzheimer’s Disease Sequencing Project (ADSP) and the Alzheimer’s Disease Neuroimaging Initiative (ADNI). We tile 11,762 genomes, a subset of the release which does not include family-based datasets (AD Cases: 4,983, age range: 50-90 years, mean age: 73.8 years). We illustrate the use of tiled data in ML classification methods to predict phenotypes. Specifically, we identify and prioritize tile variants/genetic variants that are possible genetic signatures for AD. The model shows added predictive value from variants of genes previously found to be associated with AD risk, age of onset, neurofibrillary tangle measurements, and other AD-related traits–including the APOE variant (rs429358).

## 1. Introduction

The scientific and medical community is reaching an era of affordable whole genome sequencing (WGS), opening the possibility of precision medicine for millions of individuals. As genomic datasets grow to an unprecedented scale, there is an opportunity to revisit how genomes are represented for use in clinical decision making as well as to facilitate machine learning (ML) algorithms that analyze these large-scale WGS data. There is a need for ML methods to handle the complexity and size of WGS data, which remain extremely challenging to analyze. Traditional techniques such as genome-wide association studies (GWAS) have been established for handling the analysis of genotype data, but currently very limited tools exist to analyze WGS data at a similar scale. WGS data is typically over 1000x larger than the SNP-based genotyping data used for traditional GWAS studies, making it much more computationally intensive and algorithmically challenging to analyze.

To aid in these large-scale analyses, we present *whole genome tiling*: a flexible, concise representation of whole genome sequences that supports simple and consistent names based on tile hash, ML, and annotations. The overarching goal with tiling is to provide a standard, technology independent genomic representation alongside open source ML workflows that enable the analysis of large, distributed data from WGS projects (10,000-1M+ people). We illustrate the utility of the tiling method by showing examples of ML on WGS data with a ∼10K sample size as part of the Artificial Intelligence for Alzheimer’s Disease initiative (AI4AD).

The AI4AD initiative focuses on analytic approaches for ultra-scale analysis of Alzheimer’s disease (AD) genomic and phenotypic data. This includes using ML and artificial intelligence (AI) (including deep learning), large-scale data integration, and international data harmonization to discover features associated with AD risk, diagnosis and treatment response, in massive scale genomics data. As part of this work, we represent whole genome sequences available from the Alzheimer’s Disease Sequencing Project (ADSP) and the Alzheimer’s Disease Neuroimaging Initiative (ADNI) to create a standardized and curated library of ‘tiles’ that are read into ML methods to identify AD risk and protective factors.

## 2. Materials and Methods

### 2.1 Tiling the genome

Tiling is a flexible, efficient representation of genomic data that enables fast queries and machine learning. Tiling abstracts a called genome by partitioning it into overlapping shorter segments–termed tiles. A tile is a variable length genomic sequence that is braced on each side by a 24 base (24-mer) “tag.” 24-mer tags were chosen as they were the shortest *k*-mer that allowed us to adequately tile the genome. Short tags are preferred to limit the occurrence of no-calls or genomic variants on the tags themselves. Tags that can uniquely place on the reference genome are chosen from the superset of all 24-mers. **Figure 1** explains the relationship between: tags, tag sets, tile position, tiled genomes and the tile library.

**Figure 1:**
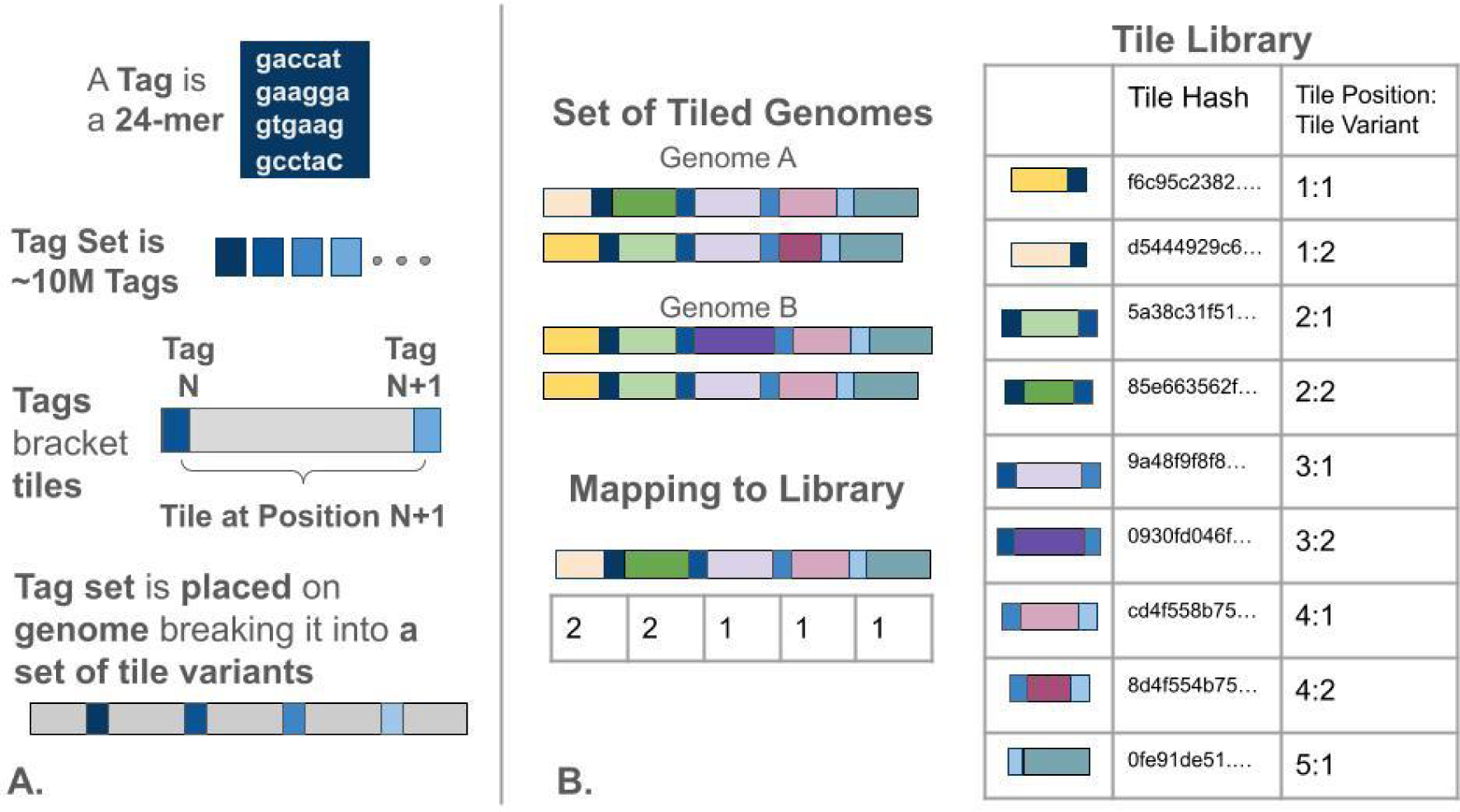
**A.** We show the relationship between a single tag (24-mer), the tag set (a fixed set of 24-mers used to partition any human genome into tiles) and the placement of tags on a tile variant. **B.** Diagram illustrating a set of 2 tiled genomes (each allele tiled separately), the tile library of tile variants that resulted from tiling those genomes and how tiled genomes are mapped to the tile library representing any genome as an array of integers.

In our implementation, a tile sequence must be at least 248 base pairs long, where each tile is labeled with a “position” that is named by the tags that brace the tile sequence. Tags are placed in the same order as they are identified in the reference genome. Tile positions are based on the placement of tags on the genome. The base-pairs between those tags including the tags themselves are then identified as a tile variant. One tile position can have multiple tile variants, one for each sequence observed at that position. Each tile variant is labeled with a hash digest (e.g. Secure Hash Algorithm 2) of the sequence it contains. When a genetic variation or no-call occurs on a tag, the tag is considered to not match the tag set and the tile is extended past the un-matched tag until a matching tag is found. Thus, tile variants can span across tile positions where the tile variant would normally end. These tiles that span multiple tile positions are known as “spanning tiles”.

Our choice of tags partitioned the GRCh37 human reference genome into 10,655,006 tiles, composed of 3.1 billion bases (an average of 314.5 bases per tile). This tag set also works well with GRCh38, with the placement of 99.8% of the tag set. We predict that changes in the tag set might be useful in the future, based on new reference genomes/pangenomes being developed. If we choose a new tag set to better represent pangenomes, the tag set would be expanded without the removal of existing tags, and we would provide tools to losslessly re-encode regions where new tags would be better for that individual. As a result, an individual tiled genome will change less than if they had to be re-encoded for a new reference genome. Annotations are added to and stored in an annotation library, which is population-wide. Since individual sequences use pointers to tile variants in the tile library, all individuals with a pointer to a tile variant in an annotated library will be annotated identically, with all tile variant and tile position annotations.

The major advantages of tiling are: 1) A set of genomes can be represented as a compact numerical matrix so that we can easily use them with standard ML and big data methods. 2) Tiled data represents the full genome, including homozygous reference calls and both phases. Therefore, we know if regions are confidently called as reference or have variants. 3) Tiling is reference and sequencing technology independent. This makes it possible to harmonize different studies that use a different reference (e.g., GrCh37 vs GrCh38), different sequencing technologies, and/or to integrate genome, exome and microarray data. Tiled data is compact and scalable. The human reference genome becomes ∼10M tiles vs 3B bases. Unlike representation based on the number of unique SNPs - which grows with the size of the cohort - tiled data will always be represented by ∼10M tile positions. Adding new genomes does not change the number of tags (or tile positions).

For an illustrative example, one can consider the 1000 Genomes cohort where there are about 18,212,589 common alleles and 45,931,977 rare alleles. To represent both alleles in the cohort with GWAS additive encoding, an analyst would have approximately 63-128 million columns of data, This is in contrast to the 20 million columns (features) in tiling. That 20 million is the same as the cohort size increases, unlike the GWAS encoding. Once cohorts grow from 100Ks to millions of genomes, the traditional encoding will not be feasible. Tiling minimizes initial data size without affecting ML or modeling approaches, facilitating efficient windowing in neural network design. Additionally, the integer representation of tile variants uses zero to represent the most common tile – allowing tiled data to be held in sparse matrices to decrease memory usage and speed numeric calculations.

### 2.2 Whole genome sequencing data

Data used in the preparation of this article were obtained from the Alzheimer’s Disease Sequencing Project (ADSP). Launched in 2012, the Alzheimer’s Disease Sequencing Project (ADSP) focuses on identifying genetic variants associated with AD by leveraging high-throughput sequencing technologies. It provides whole-genome and whole-exome sequencing data, as well as data from family-based and case-control studies. ADSP aims to discover rare and common genetic variants associated with dementia risk and progression, and their functional impacts. The project provides extensive datasets accessible to qualified researchers through application processes, making it easier to develop predictive models, identify therapeutic targets, and advance personalized medicine for AD.

A subset of the ADSP WGS data is from the Alzheimer’s Disease Neuroimaging Initiative (ADNI) database (adni.loni.usc.edu). The ADNI was launched in 2003 as a public-private partnership, led by Principal Investigator, Michael W. Weiner, MD. The primary goal of ADNI has been to test whether serial magnetic resonance imaging (MRI), positron emission tomography (PET), other biological markers, and clinical and neuropsychological assessment can be combined to measure the progression of mild cognitive impairment (MCI) and early Alzheimer’s disease (AD) (Petersen et al. 2010).

Our analysis focused on WGS available in Release 3 WGS of ADSP. The third ADSP data release occurred on October 27, 2021, and included WGS data for 16,905 individuals (Leung et al. 2023). Of that set, we analyze those genomes (14,474) that are available to both for-profit and not-for-profit institutions. We focus on the subset of 11,762 genomes which are not part of family-based datasets. The demographic summary of the data is: Male: 7,155; Female: 4,607; AD Cases: 4,983; age range: 50-90 years, mean age: 73.8 years. Accompanying the sequencing data is a basic set of phenotypes collected from each of the submitting cohorts and harmonized into the ADSP format. Currently, we focus on the AD case/control status for our prediction experiments. WGS is called using the SNP/Indel Variant Calling Pipeline and data management tool (VCPA). Variants are called using GATK haplotypecaller (Leung et al. 2023).

### 2.3. Modeling and feature selection with tiled data

Modeling genomic data is an extreme example of a *p* x *n* problem where *p*, the number of predictors (e.g., genetic variants), greatly outnumbers *n*, the number of samples (e.g., number of participants in a given study). When the dimensionality of the predictors is larger than the sample size, new or extended statistical methods and theories are needed. Recently, statisticians and ML experts have created new techniques to deal with these types of datasets that present challenges for linear modeling. While tiling reduces the number of input features, we are still analyzing a *p* of ∼20 million (tile variants/categorical variables) for an *n* in the range of 1-100 thousand (samples).

Our task when working with the ADSP tiled dataset is three-fold: (1) showcase the utility of tiling within standard ML frameworks; (2) to develop a reproducible, “base-level” model that can be compared to more advanced ML methods and models; and (3) provide a set of tile variants or genomic variants that can be passed on to other modeling efforts that include imaging and biomarker features.

In our base-level models (**Figure 2**), we aim to predict AD case-control status using tiled genomic data as input, in addition to five principal components to capture population structure, age, and sex. To find these important variants, we rely on feature selection. Feature selection (a.k.a. variable selection) allows us to identify the relevant subset of features to make available to the model. Ideally, we aim for the most parsimonious model. It is critical to limit the feature space as much as possible while avoiding the elimination of relevant features from consideration. We begin by encoding the genomic variables using Weight of Evidence (WOE) to handle the categorical nature of the predictors effectively. We filter on *p*-values based on a *p*-value threshold of 1x10^-8^ to reduce the number of features. P-values are calculated in 3 ways, using: (1) the log-likelihood ratio of a generalized linear model (GLM) with sex, age and first five principal components, with and without the encoded tile variants at that position; 2) chi-squared statistic for a difference in proportions between the encoded tile variants at that position and the AD case/control status; and (3) the *p*-value for the tile variant feature in the fitted GLM. GLM is fit with no regularization and uses a binomial family for logistic regression.

**Figure 2:**
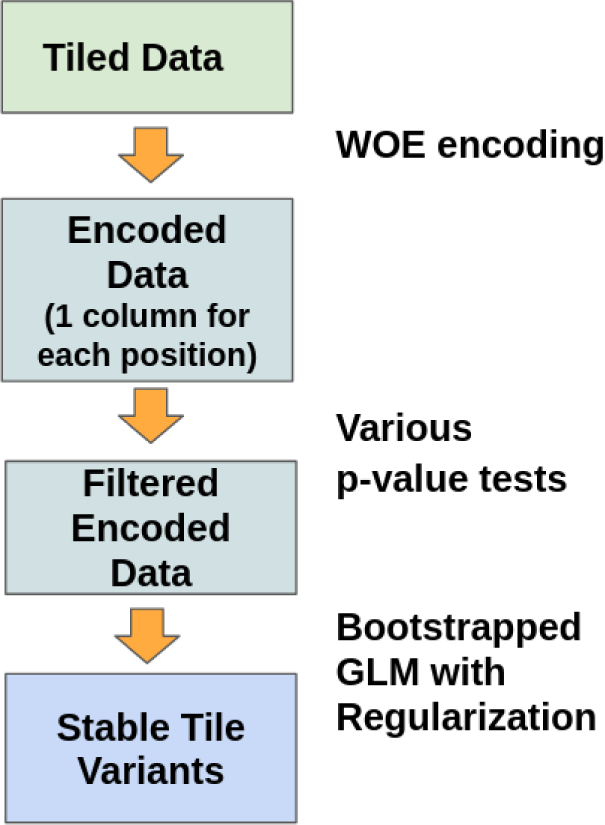
Simple diagram of modeling workflow which includes WOE encoding of the tile variants, filtering of tile variants using various significance tests, and bootstrapping GLM with regularization (adaptive lasso) to find stable tile variants. These stable tile variants are then fed into a final model for validation and comparison with models with only APOE and selected phenotype data.

Then, we bootstrap adaptive lasso models to identify stable variants. Adaptive lasso is well-suited for high-dimensional data, and helps in feature selection by assigning weights to each variable, shrinking some coefficients to zero and retaining only the most significant predictors (Zou 2006). The APOE region is known to have a strong and established effect on Alzhimer’s risk and protection. This strong effect can overshadow other genetic factors, making it harder to identify these factors and their effects. To address this, we have removed the APOE-related tile positions from our tile position feature set. We keep tile positions within TOMM40 and APOC genes since there is evidence that these may contribute to AD risk, independently of APOE. The effect of APOE4 status is modeled in a simple logistic regression model using the training set and those results are used as an offset in our main GLM model with adaptive lasso. An offset is used to account for known effects that the user does not want to estimate within the model. Essentially, the offset allows a user to include a term in the linear predictor that is not associated with any of the model’s coefficients.

To determine the stability of these significant features, we repeatedly sampled from the dataset and refitted the model to generate an ensemble of predictions. Features that appear frequently and with significant coefficients in multiple iterations are considered stable and reliable predictors. These stable features can provide insights into the underlying genetic factors associated with the model phenotype (in our example: late-onset Alzheimer’s disease). This involves repeatedly sampling the data and fitting GLM for binary classification, using cross-validation to determine the optimal regularization parameter for the adaptive lasso penalty. The process highlights the most consistent and reliable predictors across multiple iterations. Finally, we construct the final model using these stable variants, ensuring a robust, well-generalized model.

### 2.4. Weight-of-Evidence encoding

Weight of Evidence (WoE) (**Equation 1**) encoding is a powerful technique for transforming categorical variables into continuous variables, making them suitable for statistical analysis, particularly in logistic regression models. This method involves calculating the WoE for each category within a categorical variable based on the proportion of event and non-events. In our case, we consider a binary classification where our target variable is case (event) and control (non-event).

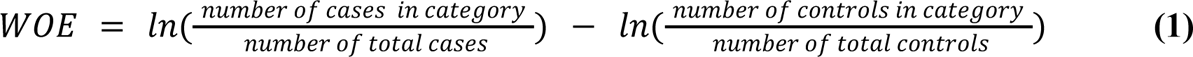

We apply an offset in Weight of Evidence (WOE) to ensure that the no-effect point is zero. When encoding that involves both tile variants and race and ethnicity phenotype, we calculate the WOE relative to the race and ethnicity category totals (not full set totals) and offset, so that the no-effect point is zero relative to the race and ethnicity phenotype. This allows for accurate comparison of tile variants across race and ethnicity categories with different initial ratios.

WoE encoding is advantageous because it often results in a monotonic relationship between the encoded variable and the target variable, improves the interpretability and predictive power of the model, and can handle missing values by assigning them a specific category. Thus tWoE encoding ensures more accurate and reliable predictions.

Specifically, in encoding tile variant data for each tile position, WOE allows for a compact encoding when compared to other encoding methods such as one-hot–which expands features from each tile position to having two features for each unique tile variant (one for heterozygous occurrence and one for homozygous occurrence). Currently, since our data is not globally phased, we encode pairs of tile variants per position (one for each allele) and order them, so that tile variant pair 2-1 and tile variant pair 1-2 are treated as the same tile variant. However, it is easy to adjust the model to allow for globally phased data by treating 2-1 and 1-2 as distinct pairs. Additionally, we can merge the given race and ethnicity phenotype with our tile variants to allow us to encode tile variants for different phenotypes differently. If phenotype does not matter, then the encoding will simply map the different categories to the same value. We show an example of this type of encoding in Table 1, which shows the differing effects of APOE4 for different ethnoracial ancestry categories in this dataset. For all ancestry categories, we see the influence of tile variants containing the APOE4 variant increasing from the heterozygous occurrence (i.e., tile variant exists on 1 allele) to homozygous occurrence (i.e., tile variant exists on both alleles). Additionally, we see the interaction of two SNPs on the same tile variant (tile variant 4 with rs769455, rs429358). The added SNP increases the WOE value relative to a tile with heterozygous rs429358 only. This effect has been noted recently in work by Le Guen et al., who found that rs769455 is associated with an increased risk of AD among individuals with the ε3/ε4 genotype.

**Table 1.** WOE encoded variants at the tile position containing rs429358, a key variant in the APOE haplotype.

| TileVar | Genomic Variant | Ethnicity | Race | Adjusted WOE |
| --- | --- | --- | --- | --- |
| 2-2 | rs429358 (hom) | Not Hispanic/Latino | Black or African American | 2.37 |
| 2-2 | rs429358 (hom) | Hispanic/Latino | Other | 2.34 |
| 2-2 | rs429358 (hom) | Not Hispanic/Latino | White | 0.55 |
| 1-2 | rs429358 (het) | Not Hispanic/Latino | Black or African American | 0.76 |
| 1-2 | rs429358 (het) | Not Hispanic/Latino | White | 0.40 |
| 1-2 | rs429358 (het) | Hispanic/Latino | Other | 1.35 |
| 1-3 | rs769455 (het) | Not Hispanic/Latino | Black or African American | 0.53 |
| 1-3 | rs769455 (het) | Hispanic/ Latino | Other | 0.48 |
| 1-4 | rs769455 (het),<br>rs429358 (het) | Not Hispanic/Latino | Black or African American | 2.45 |

## 3. Results

In this section, we illustrate the utility of the tiling method by showing results using ML on a WGS dataset as part of the Artificial Intelligence for Alzheimer’s Disease initiative (AI4AD). To simplify our analysis, we will focus on the White (Non-Hispanic/Latino) sequences, which comprise approximately 6,500 genomes.

### 3.1. Tile variants associated with Alzheimer’s disease: Statistical significance

To identify significant features within our dataset, we first calculated *p*-values for each tile position in our dataset. The *p*-values can be plotted in Manhattan-style plots similarly to those used in GWAS studies (**Figure 3**). When plotted, the *p*-values show the expected peak at the location of the APOE4 variant (rs429358).

**Figure 3:**
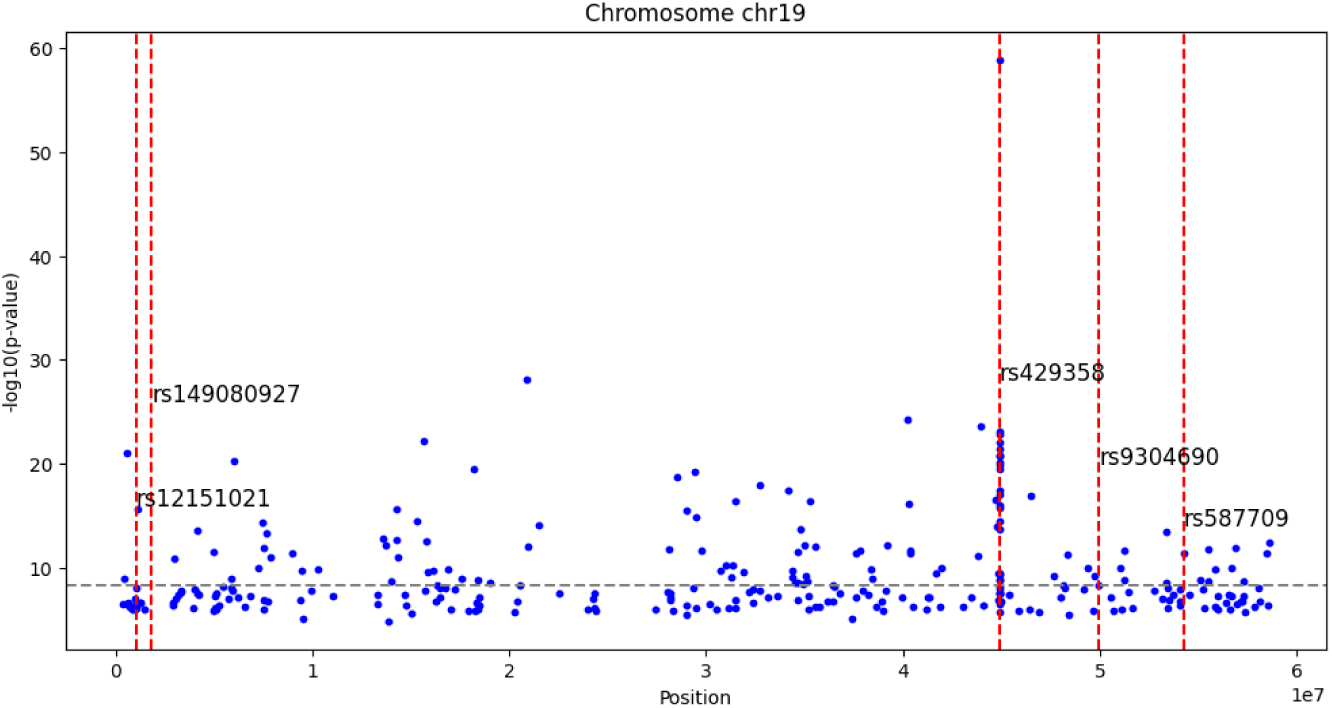
Manhattan-style plot of minimum *p*-values for encoded tile positions located on Chromosome 19. For a clearer plot, *p*-values over 1e-6 are not plotted. Gray dashed horizontal line represents the genome-wide significance threshold. The threshold has been adjusted as per the Bonferroni correction to account for the increase in the number of independent tests (set here, conservatively, to the number of tile positions). Vertical lines represent SNP positions from the ADSP AD Loci and Gene List (https://adsp.niagads.org/gvc-top-hits-list/) that occur in Chromosome 19.

Each value indicates the statistical significance of the differences observed within the respective tile. The *p*-values for the top 5 *p*-values along with tile location, the gene and recent GWAS findings associated with that gene or variant are located in **Table 2**. As expected, the tile position – and therefore the tile variant containing (rs429358) – has the lowest *p*-value. Our results identify tile variants located in genes that are consistent with those found in other GWAS studies of AD or AD-related traits (Saunders et al. 1993; Yan et al. 2021; Coltell et al. 2021; Davies et al. 2018; Richardson et al. 2022; van der Meer et al. 2020; Shadrin et al. 2021; van der Meer et al. 2021; Wang et al. 2020; Homann et al. 2022a), indicating that tiling and our model is likely effective and reliable. We filter the tile positions by a threshold of *p*-values < 1e-8. These tile positions are then passed onto the GLM model.

**Table 2:** List of top 5 *p*-values for tile variants. We list the tile location, position in the genome, genes associated with that position, and relevant GWAS studies for that gene.

| Tile location | $P$-value | Position | Gene | GWAS Studies |
| --- | --- | --- | --- | --- |
| 9553646 | 1.34E-59 | chr19:<br>44908547-44908796 | APOE | Alzheimer's disease; Alzheimer's disease or family history of Alzheimer's disease; Cerebral amyloid deposition (PET imaging) |
| 7696319 | 1.40E-41 | chr13:<br>56401049-56401298 | SPATA2P1-RN7SKP6 | Adiponectin measurement, cholesterol:total lipids ratio, high density lipoprotein cholesterol measurement, general cognitive ability |
| 9175814 | 2.67E-35 | chr18:<br>21810661-21810910 | MIB1 | Brain morphology (MOSTest) |
| 5635361 | 3.184E-32 | chr8:<br>141284277-141284527 | RP11-10J21.5;<br>SLC45A4 | Vertex-wise cortical surface area, Vertex-wise sulcal depth, Cortical thickness, Cortical surface area, Vertex-wise cortical thickness, Total PHF-tau (SNP x SNP interaction) |
| 9168826 | 1.0E-30 | chr18:<br>13498825-13499160 | LDLRAD4;<br>RP11-53B2.5 | General cognitive ability, Global cognition (Mini Mental State Examination), Total PHF-tau (SNP x SNP interaction) |

### 3.2 Tile variants associated with Alzheimer’s disease: Bootstrapping GLM models

To predict AD case-control status, we use a Generalized Linear Model (GLM) with adaptive lasso regularization. Even though cross-validation is used, the bootstrap GLM models with adaptive lasso tend towards overfitting – having sets of features that are unique to individual bootstrapped models. This is visible in the area under the curve values for the bootstrapped models, which are ∼0.94 for the training set, while the validation set AUC is ∼0.63. This can then be compared to results of the GLM trained using APOE4, Sex, Age, and PC1-PC5. In this model, APOE4, Age and PC5 are selected, yielding an AUC of 0.61.

Features that appear frequently and with significant coefficients in multiple iterations are considered stable and reliable predictors. These stable features (**Table 3**) can provide insights into underlying genetic factors associated with late-onset Alzheimer’s disease. For each stable feature, we examined the corresponding genomic regions to identify the genes in which these features reside. This step is crucial for understanding the biological relevance of the predictors. By mapping the features to specific genes, we can better infer potential roles in AD pathogenesis.

**Table 3:** List of tile variants kept in 100% of the bootstrap models. We list the tile location, position in the genome, genes associated with that position, and relevant GWAS studies for that gene.

| **Tile Variant** | **Genomic Position** | **Gene** | **GWAS studies** |
| --- | --- | --- | --- |
| 987620 | chr22: 24840698- 24841233 | SGSM1 | Global cognition (Mini Mental State Examination) |
| 2322921 | chr3: 153472368- 153472661 | LINC02006 | Cerebral amyloid deposition (PET imaging), High density lipoprotein cholesterol levels |
| 7125710 | chr12:18658599-18658863 | PIK3C2G-PLCZ1 | Alzheimer's disease |
| 7991555 | chr14: 37007797-37008178 | SLC25A21 | Total PHF-tau (SNP x SNP interaction) |
| 1879058 | chr3: 35128083-35128809 | FECHP1-KRT8P18 | Entorhinal cortical thickness |
| 7544949 | chr12: 131291294-131291864 | RPS6P20 |  |
| 3521592 | chr5: 85944260- 85944509 | PTP4A1P4 | Total PHF-tau (SNP x SNP interaction) |
| 8262259 | chr15: 21269142-21269531 | RP11-32B5.2-RP11-32B5.1 |  |
| 9553585 | chr19: 44891894-44892143 | TOMM40 | Alzheimer's disease onset, low density lipoprotein cholesterol measurement, Cerebrospinal fluid p-Tau181p:AB1-42 ratio, Hippocampal atrophy rate |
| 5170845 | chr8:19635569 -19636028 | CSGALNACT1 | High density lipoprotein cholesterol measurement, low density lipoprotein cholesterol measurement, Apolipoprotein A1 levels, Dementia in non-APOE ε4 carriers |
| 8261770 | chr15: 20344885-20345147 | CHEK2P2, HERC2P3 |  |
| 4766881 | chr7: 68339007-68339301 | RP5-945F2.3-RP5-859M6.1 |  |
| 9553660 | chr19: 44912686- 44913056 | APOC1 | Alzheimer's disease/ family history of Alzheimer's disease, Low density lipoprotein cholesterol levels, Total cholesterol levels, Middle temporal cortical thickness, Cerebral amyloid deposition (PET imaging) |

Furthermore, we cross-referenced these genomic positions with previous GWAS findings to determine whether these features have been previously associated with traits or phenotypes relevant to Alzheimer’s disease(Homann et al. 2022b; Yan et al. 2021; Sinnott-Armstrong et al. 2021; Kunkle et al. 2021; Wang et al. 2020; Guo et al. 2019; Liu et al. 2018; Wojcik et al. 2019; Lo et al. 2019; Harper et al. 2022; Graham et al. 2021; Richardson et al. 2022; Yan et al. 2021; Park et al. 2021; Schwartzentruber et al. 2021). The convergence of our model’s features with established GWAS associations supports our model’s performance and its ability to capture meaningful genetic signals. Using only stable features that occur in 75% of bootstrapped models, we fit a non-regularized logistic regression which performs similarly to the bootstrap models, AUC: 0.62.

## 4. Discussion

Genomic tiling, introduced in this paper, is a method to break down the genome into smaller, manageable segments, known as tiles - without loss of information. The tiling encoding is lossless because it preserves every confidently called base (including confidently called homozygous reference bases). Genomes can be in any format or use any genomic reference — consistent with recommendations for use of such data in clinical settings (Lubin et al. 2017)— or could be telomere-to-telomere haplotype resolved assemblies (Peters et al. 2012; Marbl, 2024) Since short-read sequencing typically leaves genomes unphased, tiled genomes can also be phased and imputed prior to tiling. Each tile represents a specific region of the genome and can include multiple genetic variants. This approach allows for a comprehensive and systematic analysis of the genetic data, enabling researchers to capture detailed information about the genetic architecture of complex traits, including variants that would be missed in conventional GWAS analyses.

We presented machine learning (ML) models that work well with tiled genomic data, illustrating that tiling can handle large-scale, high-dimensional datasets effectively. ML models can uncover associations within the data that might be missed by traditional mass-univariate statistical methods. By evaluating the performance of these models on tiled genomic data, researchers can ensure that the models are robust, reliable, and capable of providing valuable insights into the genetic underpinnings of diseases, such as Alzheimer’s disease. This, in turn, facilitates more accurate predictions, better understanding of disease mechanisms, and the potential identification of novel therapeutic targets.

Tiling is particularly advantageous for AD modeling as it can capture a comprehensive and detailed genetic landscape, while representing the genome concisely. Heritability estimates for AD, which represents the proportion of phenotypic variance attributable to genetic factors, vary between 38% and 79% (Gatz et al. 2006). This considerable heritability suggests that a significant portion of heritability remains unexplained by current genetic models. By using tiled genomes, researchers can systematically represent and analyze the genetic variants across entire genomes, including common and rare variants identified by genome-wide association studies (GWAS) and whole-genome sequencing. This tiling approach not only enhances the resolution of genetic data but also simplifies the representation and management of the genome, allowing for a more precise identification of the genetic factors contributing to AD, clarifying the complex genetic architecture of the disease.

Our model shows added predictive value for variants in genes previously to be associated with AD risk, and with paired helical filament tau measurements, neurofibrillary tangles measurements, and hippocampal atrophy.

## 6. Acknowledgments

This work was funded in part by NIA grant U01 AG068057 to the AI4AD Consortium. Data collection for ADNI was also supported by the NIA and a combination of public and private funders. ADSP data collection was supported by the NIA (see supplementary materials for detailed acknowledgements for NIAGADS, ADSP and ADNI.)

## Data Availability

All data produced in the present study to be obtained from NIAGADS (https://www.niagads.org/) DOI in process.

## Supplemental Materials

### S1. Detailed Acknowledgements

Data for this study were prepared, archived, and distributed by the National Institute on Aging Alzheimer’s Disease Data Storage Site (NIAGADS) at the University of Pennsylvania (U24-AG041689), funded by the National Institute on Aging. See the following link https://dss.niagads.org/datasets/ng00067/#dataset-acknowledgement for additional acknowledgements for ADSP data.

Data collection and sharing for this project was funded by the Alzheimer’s Disease Neuroimaging Initiative (ADNI) (National Institutes of Health Grant U01 AG024904) and DOD ADNI (Department of Defense award number W81XWH-12-2-0012). See this link for additional information and acknowledgments for the ADNI data: https://adni.loni.usc.edu/data-samples/adni-data

### S2. Supplemental Methods

#### S2.1. Code for tiling the genome

Preparing a set of samples is a multi-step process, run as a CWL workflow on the Arvados platform. First, each sample is filtered using a genotype quality (GQ) of 30 or higher, converted from GVCF to a matched set of VCF and BED coverage files, then converted to a pair of FASTA files using the bedtools consensus utility. The resulting FASTA dataset is split into batches of 100 samples for tiling. Each tiling job reads a batch of 100 FASTA genomes, locates all of the 24-mer tags, assigns a number to each distinct variant (i.e., the sequence between tags), and outputs a tile library containing the sequence data for each distinct variant found in the batch, along with each sample’s tile representation: an array with two variant numbers for each 24-mer tag. The next step is to collate the 100-sample batches into 214 “slices,” where each slice contains the sample data for the entire population in one 50K-tag portion of the genome. This batch/slice approach is needed to process a large dataset without requiring a correspondingly large amount of RAM.

The tiling process itself is quite efficient. The FASTA data is read, each base pair is converted to a 2-bit representation, and bitwise operators (x ← x << 2 | b) are used to produce a 48-bit integer representing the last 24 bases. This is looked up in a hash table of 2-bit-encoded tags. When a tag is found, the blake2b hash of the tile variant sequence (between this tag and the previous tag) is compared to a list of known variants for the same starting tag, and the sequence is written to disk if it is new. The Go runtime’s concurrency model allows efficient use of multiple CPU cores to tile a batch of genomes. Typically a given variant will be numbered differently when it appears in multiple batches; this is resolved in the “slice” stage where equivalent variants in different batches are identified by their blake2b hashes, and renumbered according to their overall frequency in the merged dataset.

As one would expect, the sequence of tags found in a sample genome is not identical to the sequence of tags in the reference genome. Some tags appear out-of-order, some appear multiple times, and some do not appear at all. Currently, these outlier tags are simply skipped: this means, for example, that a variant of tile 100 might start with tag 100, end with tag 101, and contain tag 123 in between those two; and a sample referencing such a variant would therefore not have a variant for tile 123, but might have a spanning tile beginning with tag 122 and ending with tag 124. This strategy results in some tile variants being longer than they would if every tag were used, but it ensures that every sample’s tiling can be expressed as an array with two variant numbers per tile position. The vast majority of tiles are unaffected by these issues, partly because the input sequences themselves were constructed by aligning short reads to a reference sequence.

To annotate tiles, HGVS variants are generated from the tiled data, then looked up in annotation databases using available tools. To generate the HGVS variants, the GRCh38 reference genome is tiled, and each tile variant in the sample set is compared to the tile variant found in the reference with the same starting tag. The process starts with a generic diff algorithm; the result is adjusted to match HGVS guidelines, an offset corresponding to the location of the start tag in the reference genome is added, and the diff is formatted as an HGVS ID. The entire set of annotations is output in the form of a VCF file: each line has a single HGVS variant and a list of the tile variant IDs in which that HGVS variant was found. The VCF files are then annotated using SnpEff, an open source tool that performs annotation on variants and predicts their effects on genes.

#### S2.2. Pythonic tiling representation for machine learning

The tiled genomes are saved as numpy-encoded matrices to facilitate easier use and management. Each matrix comprises one row per genome and a pair of columns per tag or tile position, corresponding to each allele. The elements within the matrix are integers that denote various states: -1 signifies a “low quality” tile variant with no-calls, 0 indicates the absence of a tag for the tile (meaning this genome section is covered by a spanning tile in an earlier column), 1 represents the most common high-quality variant of the tile in the dataset, 2 signifies the second most common, and so forth. For efficient loading, these numpy matrices are divided into manageable chunks. Notably, tile variants can span multiple tile positions when a tag is not found, and these are referred to as spanning tiles.

#### S2.3. WOE encoding

When encoding, each tile position is treated separately and all unique tile variant pairs at that location are encoded as a set of categorical variables. To effectively utilize the Weight of Evidence (WoE) encoding for categorical variables in predictive modeling while avoiding data leakage, a structured approach is followed using Python and the category_encoders library. Initially, the data set of tiled genomes is split into training and validation sets, ensuring that the validation data remains unseen during the training phase. The WoE encoder, specifically designed for categorical variables, is then applied solely to the training set. This step involves fitting the encoder to the training data, transforming the categorical variables into a numerical format that reflects their predictive power relative to the target variable. Cross-validation is performed using the training set to evaluate the model’s performance, ensuring it generalizes well and prevents overfitting. After cross-validation, the validation set is transformed using the previously fitted WoE encoder, maintaining consistency in data processing. This method ensures that feature encoding and model training processes are confined to the training data, preventing any leakage of information from the validation set and providing an accurate assessment of the model’s predictive capabilities.

#### S2.4. High-quality tile variants

Before encoding or calculating *p*-values, the tiled data is restricted to tile positions of high quality. High quality tile positions are defined as those having 90% or more high quality tiles at that position. High quality tiles are defined as having no uncalled bases. Imputation and phasing increases the number of quality tile positions and will be included in tiling efforts.

#### S2.5. Unsupervised learning - Principal component analysis

To avoid capturing the noise generated by sequencing artifacts, we ran PCA only on tile positions where 98% of tile variants were well-sequenced. The numpy arrays were then encoded using a one-hot encoding scheme. The resulting matrix was used as input for principal component analysis, implemented by scikit-learn (Pedregosa et al. 2011), and projected onto the first five principal components.

#### S2.6. Supervised learning - GLM with adaptive lasso

Using the *glmnet* package in R (Friedman et al. 2019), one can perform binomial regression with an adaptive lasso penalty to account for variable selection and shrinkage in a robust manner.

Generalized Linear Models (GLMs) are a flexible generalization of ordinary linear regression that allows for the dependent variable to have a non-normal distribution. The binomial family of GLMs is particularly suited for binary outcomes, making it ideal for classification problems where the response variable is dichotomous, such as the presence or absence of a disease.

The adaptive lasso is an enhancement of the traditional lasso (Least Absolute Shrinkage and Selection Operator) technique. It assigns different penalties to different coefficients, allowing for more flexible and accurate model fitting by shrinking some coefficients more than others based on their importance. To implement this, weights can be computed based on the initial estimates of coefficients, often derived from a ridge regression model. This adaptive approach can improve the model’s ability to select the most relevant variables while reducing the impact of less important ones, enhancing the interpretability and predictive performance of the model (Zou 2006).

To account for the known significant effects of variants of APOE, as defined by two SNPs, rs429358 and rs7412, an offset can be included in the regression model. This offset adjusts the baseline model to reflect the known effect of the APOE genotype, thus allowing the model to focus on uncovering additional associations while appropriately accounting for the APOE phenotype’s impact. This offset is calculated by developing a simple non-regularized GLM trained on APOE status as given in the ADSP phenotypes.

Bootstrapping a model with replacement is a powerful statistical technique used to estimate the accuracy and variability of a predictive model. In our models, we use bootstrapping to determine stable variants in our model. This method involves repeatedly sampling from the original dataset with replacement to create multiple new datasets, each known as a bootstrap sample, and running the model on that new dataset.

### S3. Tiling with Public Data

#### S3.1. Public Whole Genome Data

Along with the ADSP data, we have tiled a group of publicly available genomes from Harvard Personal Genome Project (Ball et al. 2014), Personal Genome Project Canada (Reuter et al. 2018), 1000 Genomes Project (The 1000 Genomes Project Consortium 2015) and the Simons Diversity Project (Mallick et al. 2016). This readily available public data allows researchers to immediately start working with tiled data and also provide data alongside their models and results.

**Table S1:** List of public data that we have tiled. This illustrates the ability for tiling to enable analysis between different references, variant calling pipelines and formats without having to recall your previously sequenced data.

| **Project** | **# of Genomes** | **Data Format** | **Sequencing Technology** | **Variant Caller** | **Genomic Reference** |
| --- | --- | --- | --- | --- | --- |
| Harvard PGP | 365 | TSV | CGI | CGAPipeline_2.2.0 .26 | GrCh37 |
| Harvard PGP | 96 | VCF+BED | Illumina | FreeBayes & VarScan | hg19 |
| PGP Canada | 56 | GVCF | Illumina | IsaacVariantCaller | hg19 |
| UK PGP | 89 | GVCF | Illumina | GATK - HaplotypeCaller | GrCh38 |
| 1000 Genomes | 433 | TSV | CGI | CGAPipeline_2.2.0 .26 | GrCh37 |
| 1000 Genomes | 2220 | GVCF | Illumina | GATK - HaplotypeCaller | GrCh38 |
| Simons Diversity | 276 | VCF with all positions | Illumina | GATK - UnifiedGenotyper | GrCh37 |

#### S3.2. PCA Across Public Data

A PCA analysis was calculated on the public data set illustrating that tile data can facilitate working across different references, variant calling pipelines and different formats. As expected, tiled data clusters by geographic ancestry in a similar pattern as found in previous studies working with SNPs.

**Figure S1:**
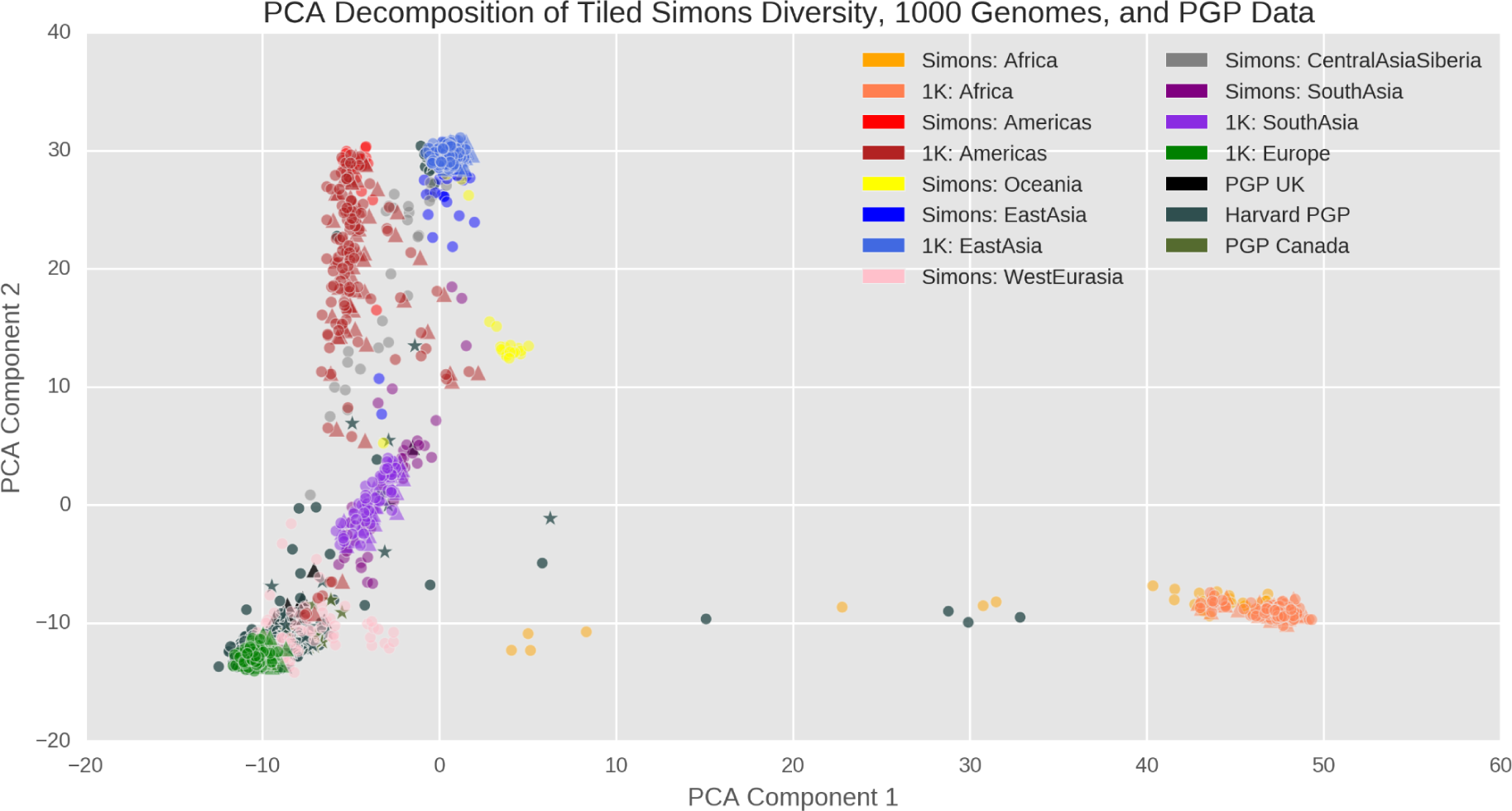
Projection of well sequenced positions in autosomal chromosomes along the first two principal components for XX whole genome sequences. Note: This figure contains a subset of 1K GrCh38 genomes to make the plot easier to read. Color represents geographic ancestry while shape represents reference genome used when sequencing: stars are hg19, circles are GrCh37, and triangles are GrCh36. Some PGP sequences come from mixed geographic ancestry as indicated by mapping between geographic ancestry clusters.

### S4. Extended AD Results

#### S4.1 Extended results for tile variants associated with Alzheimer’s disease: Statistical significance

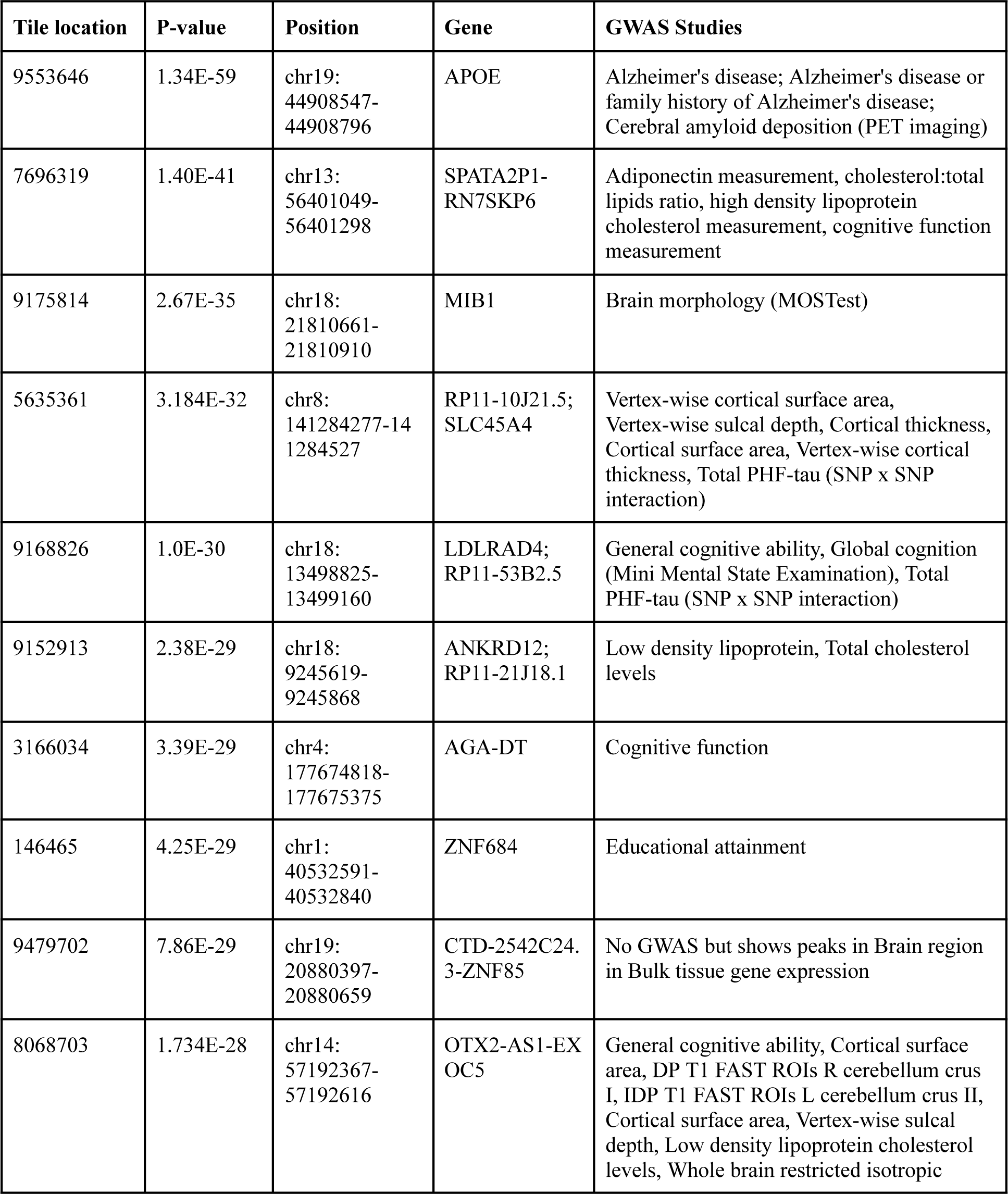
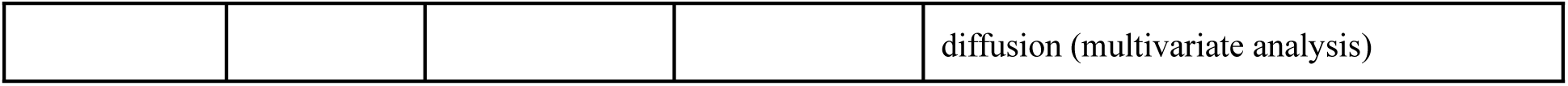

*S4.1 Extended results for tile variants associated with Alzheimer's disease: Statistical significance*
| Tile location | P-value | Position | Gene | GWAS Studies |
| --- | --- | --- | --- | --- |
| 9553646 | 1.34E-59 | chr19:<br>44908547-44908796 | APOE | Alzheimer's disease; Alzheimer's disease or family history of Alzheimer's disease; Cerebral amyloid deposition (PET imaging) |
| 7696319 | 1.40E-41 | chr13:<br>56401049-56401298 | SPATA2P1-RN7SKP6 | Adiponectin measurement, cholesterol:total lipids ratio, high density lipoprotein cholesterol measurement, cognitive function measurement |
| 9175814 | 2.67E-35 | chr18:<br>21810661-21810910 | MIB1 | Brain morphology (MOSTest) |
| 5635361 | 3.184E-32 | chr8:<br>141284277-14<br>1284527 | RP11-10J21.5;<br>SLC45A4 | Vertex-wise cortical surface area, Vertex-wise sulcal depth, Cortical thickness, Cortical surface area, Vertex-wise cortical thickness, Total PHF-tau (SNP x SNP interaction) |
| 9168826 | 1.0E-30 | chr18:<br>13498825-13499160 | LDLRAD4;<br>RP11-53B2.5 | General cognitive ability, Global cognition (Mini Mental State Examination), Total PHF-tau (SNP x SNP interaction) |
| 9152913 | 2.38E-29 | chr18:<br>9245619-9245868 | ANKRD12;<br>RP11-21J18.1 | Low density lipoprotein, Total cholesterol levels |
| 3166034 | 3.39E-29 | chr4:<br>177674818-177675375 | AGA-DT | Cognitive function |
| 146465 | 4.25E-29 | chr1:<br>40532591-40532840 | ZNF684 | Educational attainment |
| 9479702 | 7.86E-29 | chr19:<br>20880397-20880659 | CTD-2542C24.<br>3-ZNF85 | No GWAS but shows peaks in Brain region in Bulk tissue gene expression |
| 8068703 | 1.734E-28 | chr14:<br>57192367-57192616 | OTX2-AS1-EX<br>OC5 | General cognitive ability, Cortical surface area, DP T1 FAST ROIs R cerebellum crus I, IDP T1 FAST ROIs L cerebellum crus II, Cortical surface area, Vertex-wise sulcal depth, Low density lipoprotein cholesterol levels, Whole brain restricted isotropic diffusion (multivariate analysis) |

#### S4.2 Extended results for tile variants associated with Alzheimer’s disease: Bootstrapping GLM models

**Figure S2:**
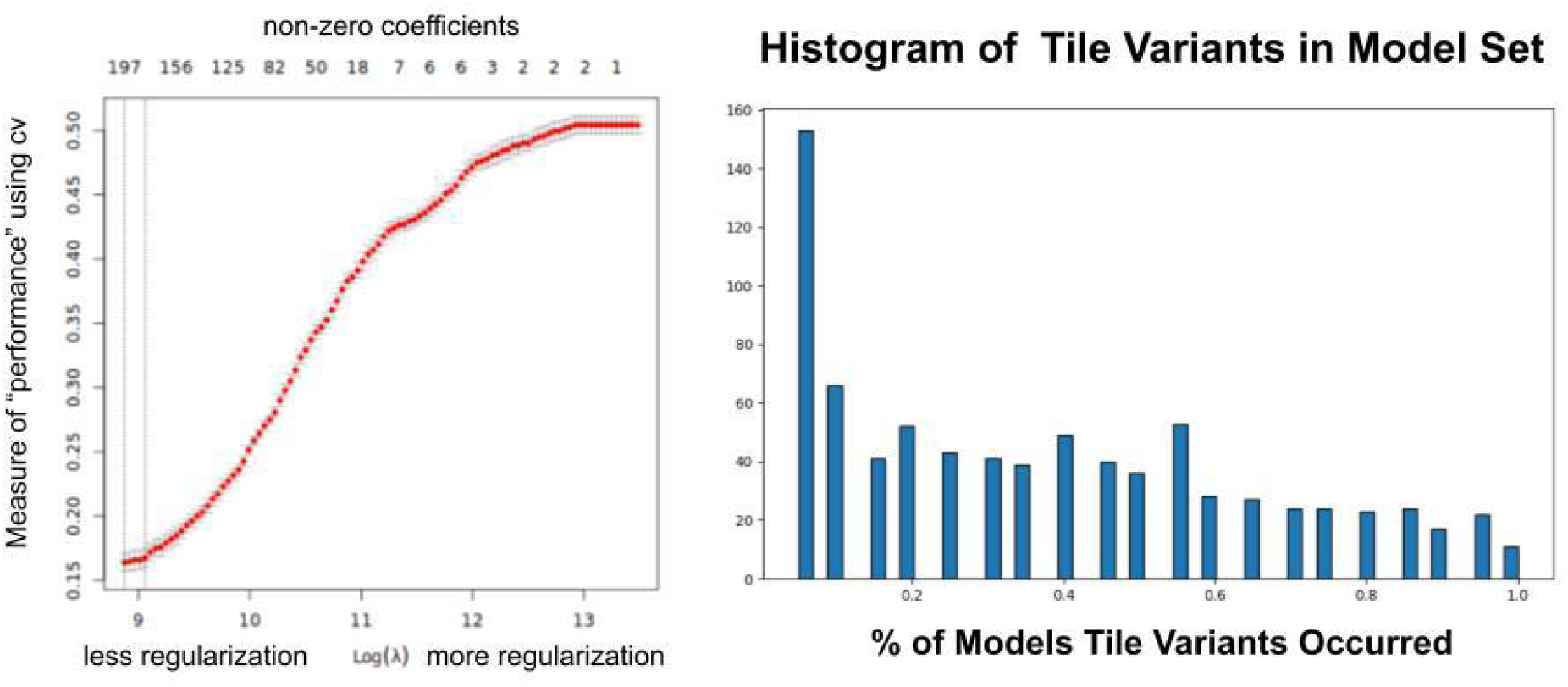
(Left) Plot of tuning regularization for GLM using cross-validation. Top x-axis shows the number of non-zero components and the bottom lambda illustrates the amount of regularization. Y-axis shows the misclassification rate for the GLM at that level of regularization. (Right) Histogram plot of number of features per fraction existing in bootstrap models.

